# Online Epileptic Seizure Detection in Long-term iEEG Recordings Using Mixed-signal Neuromorphic Circuits

**DOI:** 10.1101/2024.06.13.24308876

**Authors:** Olympia Gallou, Jim Bartels, Saptarshi Ghosh, Kaspar Schindler, Johannes Sarnthein, Giacomo Indiveri

**Affiliations:** Institute of Neuroinformatics, University of Zurich and ETH Zurich, Zurich, Switzerland; Nano Sensing Unit, Tokyo Institute of Technology, Yokohama, 226-8503, Japan; Department of Neurology, Inselspital University Hospital Bern, Bern, SwitzerlandS; Department of Neurosurgery, University Hospital Zürich, University of Zürich, Zürich, Switzerland

**Keywords:** EEG, seizure detection, event-based processing, spiking neural networks, neuromorphic processor, edge computing

## Abstract

Seizure detection stands as a critical aspect of epilepsy management, which requires continuous monitoring to improve patient care. However, existing monitoring systems face challenges in providing reliable, long-term, portable solutions due to the computational expense and power demands of continuous processing and data transmission. Edge computing offers a viable solution by enabling efficient processing locally, close to the sensors and without having to transmit the sensory signals to remote computing platforms. In this work, we present a mixed-signal hardware implementation of a biologically realistic Spiking Neural Network (SNN) for always-on monitoring with on-line seizure detection. We validated the hardware system with wideband Electroencephalography (EEG) signal recordings with over 122 continuous hours of data, without pre-filtering. The network was tested with a cohort of 5 patients and a total number of 22 seizures including generalized and focal onsets. Our system effectively captures spatiotemporal features based on synchronized multichannel intracranial EEG activity, achieving 100% sensitivity across all patients and near zero false alarms. Remarkably, inference across patients required only calibrating the parameters of the network’s output layer on a single recorded seizure from the patient.

## I. Introduction

Epilepsy is one of the most prevalent neurological disorders that affects millions of people worldwide, with an estimated 5 million individuals diagnosed each year [1]. Approximately one-third of patients do not respond to existing pharmacological treatments and continue to suffer from uncontrolled seizures [2]. These recurrent episodes can significantly influence the quality of life of patients, increase the risk of accidents, and even lead to Sudden Unexpected Death in Epilepsy (SUDEP) [3].

In clinical practice, EEG and video surveillance remain the gold standard diagnostic tools. Drug-resistant patients may need to undergo invasive channel implantation when routine EEG findings are insufficient for surgical planning. In Epilepsy Monitoring Unit (EMU), patients need to stay restricted for several days connected to bulky machinery with many wires. Both invasive and noninvasive systems require significant hospital resource investments, extensive expertise, and time for manual EEG annotation. In addition, routine practice involves patients or caregivers filling out seizure diaries, which can, however, lead to misreporting and inaccurate records [4]. These challenges underscore the need for more efficient and patient-friendly embedded monitoring solutions. Recently, the market for remote epilepsy monitoring devices has experienced substantial growth. The development of wearable and implantable technologies addresses many limitations associated with traditional restrictive monitoring systems, including the physical constraints of wired EEG setups, accessibility, and the high costs associated with prolonged hospital stays. In general, the design of such battery-powered systems should be optimized in resource-controlled environments, where size, algorithmic complexity, and energy play a crucial role. State-of-the-art EEG devices [5]–[8] are equipped with various sensors that measure physiological signals such as accelerometry, heart rate variability, electrodermal or Electromyography (EMG) activity, while integrated Machine Learning has opened a new avenue for non-invasive real-time monitoring [9]–[11].

In the literature, most of the reported seizure detection devices are still in the early stages of development. Currently, there are only a few Food and Drug Administration (FDA) or EU-cleared devices in class II; these are capable of detecting Generalized Tonic-Clonic Seizure (GTCS) in real time with non-EEG modalities [8]–[11]. There are EEG-based devices that detect specific types of seizures, including focal seizures [12] or absence seizures [13]. Two primary challenges are associated with high computational demands that reduce battery life and sub-optimal performances characterized by high False-Alarm-Rate (FAR). The need for frequent battery recharges disrupts continuous monitoring and adds a layer of inconvenience and stress to the patient. In addition, elevated FAR can lead to desensitization and reduced compliance.

Event-based neuromorphic devices have emerged as promising candidates for enabling energy-efficient biosignal monitoring and computation at the edge [14], [15] (Fig. 1). In particular, SNNs implemented in mixed-signal analog/digital neuromorphic processors have already been reported to perform spatiotemporal tasks that involve noisy electrophysiological signals efficiently [16]–[19]. In [20], an ultra-low-power Complementary Metal-Oxide-Semiconductor (CMOS)-based seizure detection approach is applied to in vitro Local Field Potential (LFP) signals, but shows very limited robustness against interictal periods carrying artifacts.

**Fig. 1:**
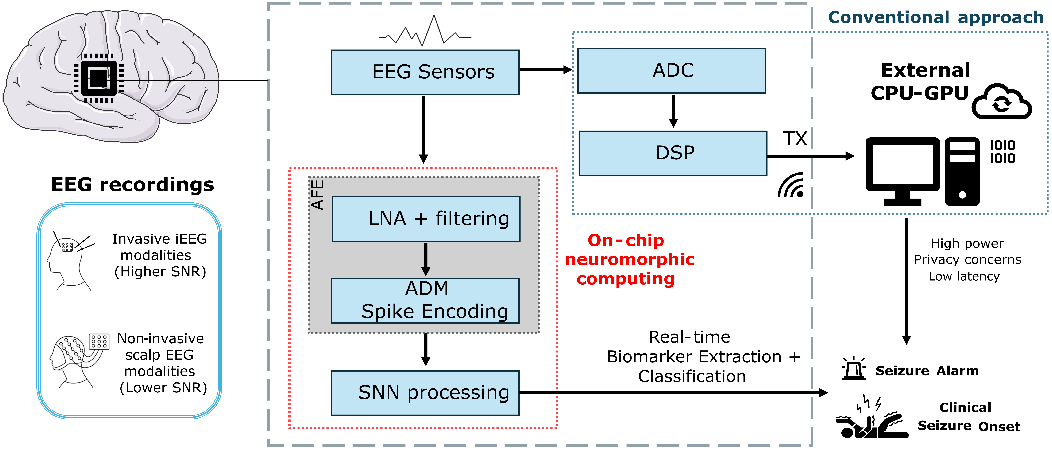
Conventional and alternative neuromorphic System-On-Chip (SoC) approaches for EEG-based seizure detection.

In this paper, we present and validate a robust spike-based hardware setup that enables long-term seizure monitoring from intracranial EEG signals using a low-power mixed-signal neuromorphic processor, with very low sensitivity to artifacts and false-alarm rates.

## II. Materials and methods

### A. Delta-modulation scheme and adaptive encoding

As the neuromorphic chip expects inputs in the form of asynchronous events, the EEG signals in the input data streams must first be converted to an event-based representation. One of the most prevalent interfacing devices used for this purpose is the Asynchronous Delta Modulator (ADM) circuit [21]. The ADM operates by encoding the changes or “delta” in the input signal asynchronously, generating events whenever the signal crosses predefined delta-thresholds. If the amplitude change in the continuous time signal since the previous event increases by a delta threshold, the ADM produces an event labeled *“UP”*, and if the signal decreases by a delta threshold the circuit produces a *“DOWN”* event. A common challenge is to establish the value of the delta threshold parameter in a way that does not significantly affect biomarker detection performance based on different noise levels: if the delta parameter is too small, the ADM will produce many events, increasing bandwidth usage and power consumption. If it is set too high, the ADM will produce too few events, and loose relevant information in the original signal. Previous applications relied on baseline-heuristics for calibrating the ADM circuits [16], [19], [22].

Recently, a novel adaptive thresholding circuit for the ADM was introduced, ideal for always-on biomedical signal processing tasks, where the encoding threshold parameter adaptively changes in real time based on the amplitude of the incoming signals [23]. This adaptive ADM circuit can dynamically adjust the delta threshold parameter to amplitude changes that may occur during long recording sessions and minimize the generation of events in background activity while retaining the information related to large anomalous fluctuations. In this work we made a hardware-aware behavioral simulation of the circuit, employing an equivalent structure to reproduce its features for implementing such adaptive thresholding feature in our long-recording data-set sessions. To reduce the computational cost of the software simulation, the adaptive delta threshold values were calculated only during a calibration period of 10 s for each channel at the start of the first interictal recording session. After this calibration phase, the delta threshold voltage values were kept fixed for each signal and the subsequent hour segments for each patient. The other ADM free parameter is its refractory period [21]. In our simulation we set this to 10 μs. The original event-based Intracranial Electroencephalography (iEEG) signal streams, which encapsulate the temporal dynamics of the brain’s electrical activity were therefore converted to a compressed stream of events “UP” and “DN” that was then transmitted to the neuromorphic spiking neural network chip.

### B. Mixed-signal neuromorphic processor

The SNN was configured on the mixed-signal Dynamic Neuromorphic Asynchronous Processor (DYNAP)-SE neuromorphic processor [24] as a prototype of an “always on” spike-based real-time seizure detection system. Each chips comprises 1024 Adaptive Exponential Leaky Integrate-and-fire (AdEx I&F) neurons, with 64 input synapse circuits each. The prototype SNN was designed to be scalable and employ a limited number of neurons (up to 106 neurons) on a single chip, for a maximum number of input EEG channels of 60, due to fan-in hardware constraints. As the synapse and neuron circuits are analog, the chip is affected by device mismatch, which can however be mitigated, using population coding and other bio-inspired processing strategies [25]. In this work, only the neuronal and synaptic parameters of two cores on the chip had to be tuned, to achieve the desired network behavior. In addition, we configured the synapse circuits to implement excitatory fast-α-Amino-3-hydroxy-5-methyl-4-isoxazolepropionic Acid (AMPA), slow-*N*-Methyl-d-aspartate (NMDA), and inhibitory shunting-γ-Aminobutanoic Acid (GABA)-A and subtractive-GABA-B type dynamics.

### C. The SNN architecture

The hardware SNN was designed by following strategies that could help overcome the device mismatch and the noisy nature of subthreshold analog circuits, such as ensemble averaging and feedforward inhibition. The architecture is a shallow network which consists of interconnected neuron ensembles, organized into two simple successive feedforward layers as described in Fig. 2, where only a subset of 12 channels is depicted, for sake of clarity. The encoded input channels are grouped into triplets that simultaneously stimulate the adjacent excitatory and inhibitory neuron ensembles of the hidden layer with feed-forward AMPA connections. Each triplet represents a subset of encoded input EEG channels whose grouping can vary depending on the number of inputs. Each hidden neuronal ensemble comprises five neurons. The inhibitory clusters send inhibitory connections to all the hidden groups of neurons except the one that was excited from the same input triplet. From the hidden layer to the output, all hidden groups project NMDA excitatory connections to an additional inhibitory cluster, and excitatory AMPA connections to the read-out neuron. The cluster of inhibitory interneurons with slower inhibitory connections in the second layer plays a normalizing regulatory role, curtailing overexcitation that could otherwise lead to mis-classification. The weights and time constants of the neuron populations in the hidden layer were uniform and tuned based on two target seizure events with focal (10 seconds) and generalized onset (40 seconds). After this tuning process, the parameters were kept fixed for all subject recordings in the hidden layer. The output layer (weights and time constants) were further fine-tuned, for each subject, based on one single seizure onset from their corresponding data.

**Fig. 2:**
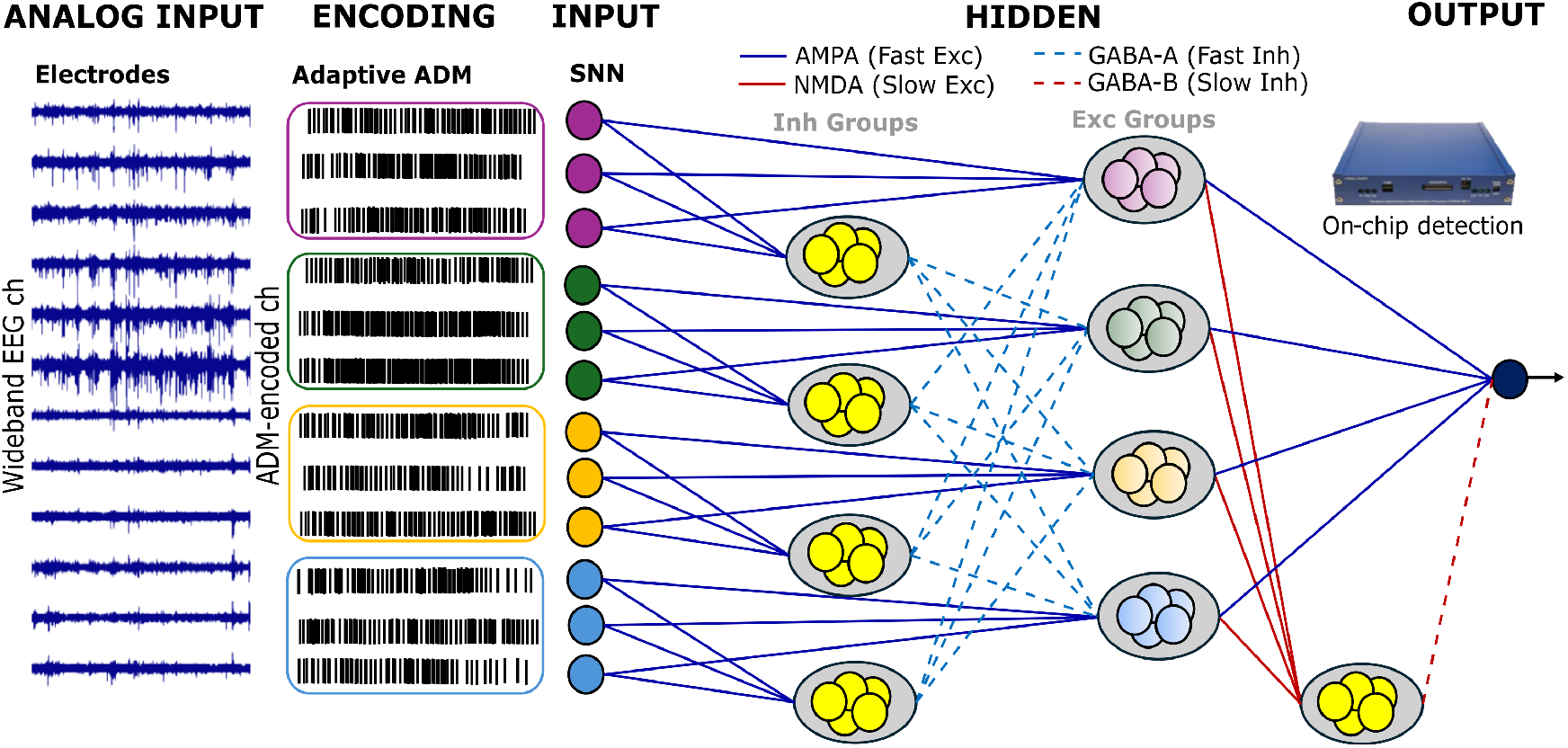
Pipeline for spike-based electrographic seizure detection with a full-SNN architecture implemented on a neuromorphic processor.

### D. Human Long-term iEEG dataset

We tested our network on continuous long-term recordings from the publicly available SWEC-ETHZ iEEG database [26]. The database includes hourly segmented recordings from drug-resistant patients who were monitored intracranially using grid, strip, or depth electrodes. After converting the analog signals to 16-bit digital format, median referencing and digital bandpass filtering between 0.5 Hz and 120 Hz, were applied. Expert annotations (onset and offset of the seizure) are provided for each ictal hour. We tested recordings of 5 patients with up to 60 input channels and 22 seizures with generalized and focal onsets. The analysis involves continuous event-based processing, allowing observation and detection of epileptic seizure events in their entirety.

## III. Results

### A. The SNN behavior

Figure 3 shows the average firing rates of the ADM encoded input channels (top plot) and hidden excitatory and inhibitory neuron groups (bottom plot) during one hour. As shown, feedforward inhibition leads to a reduced firing rate in the hidden layer, while the read-out neuron in the output layer responds to sustained synchronized activity with a rapid increase in the firing rate shortly after the marked seizure onset. Interictal periods are characterized by near-zero firing rates in all patients, highlighting the output neuron’s inactivity or minimal activity in the absence of seizures. Based on this behavior, the estimated average power consumption of the chip, averaged across all configurations adapted to the varying number of channels for each patient is P=12.48 μW (see [27] for the details on the formula used to estimate the power consumption of each circuit component in the chip).

**Fig. 3:**
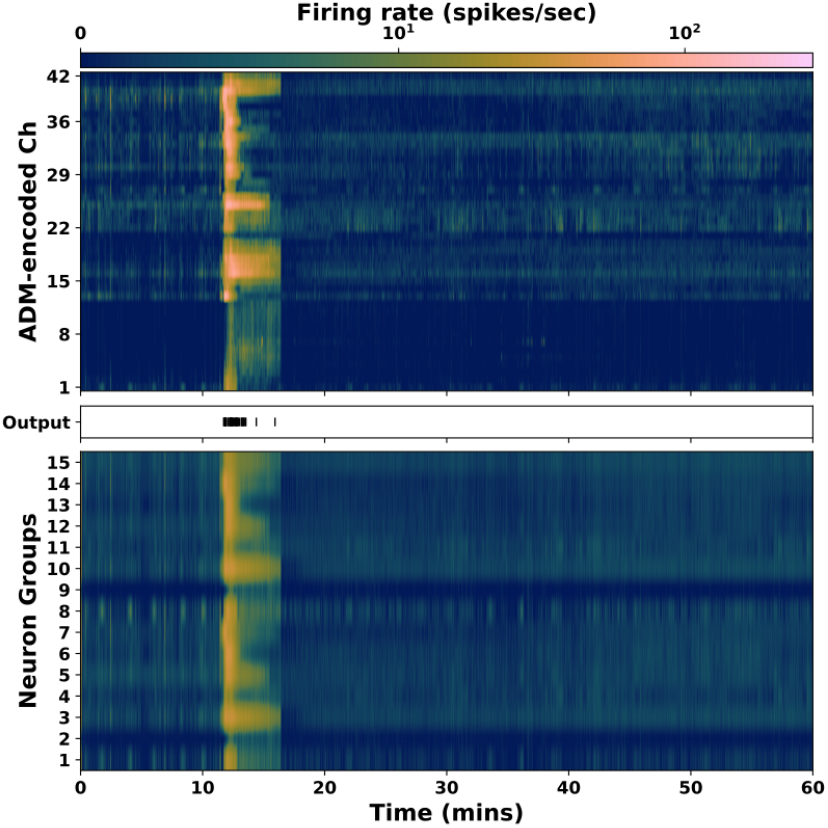
Firing rates of the ADM encoded input channels, SNN neuron groups and output during one hour of recording.

### B. Synchronization scores and correlation matrices

The degree of synchronization in different stages, from the original EEG to the encoded ADM input and hidden activity of the SNN, was quantified using group-specific correlation matrices and averaged synchronization scores (see in Fig. 4). The correlation matrices illustrate the similarity between the amplitude envelopes of different original EEG channel sets, while the synchronization scores, calculated with a coincidence window of 100 ms, indicate the ratio of observed coincidences to those expected by chance, providing a normalized measure of synchrony. These results indicate a consistent trend across both measures, suggesting that the amplitude synchronization of the EEG signals is mirrored in the temporal synchrony of the spike trains during an ictal period. Higher synchronization scores from ADM-encoded inputs correlate with higher CIs in the SNN hidden layers, demonstrating that the network effectively captures the relevant synchronization features to accurately detect seizure activity.

**Fig. 4:**
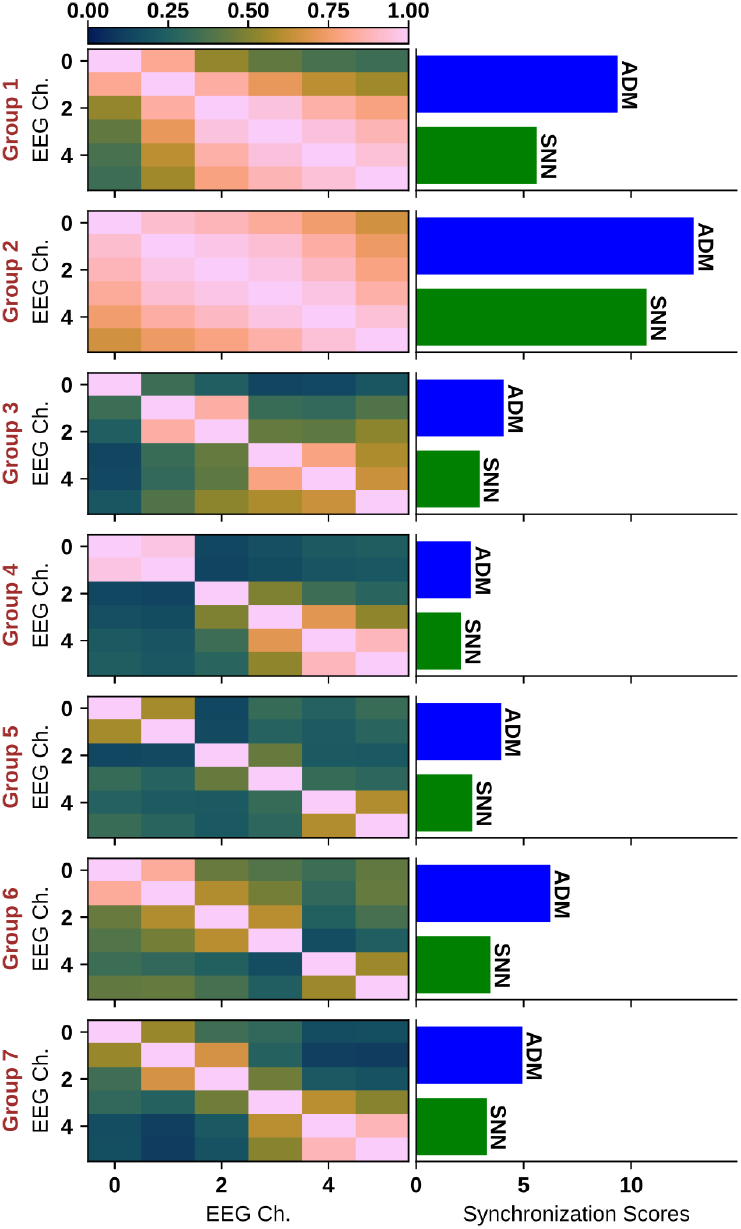
Averaged Synchronization scores within groups based on Coincidence Index (CI).

### C. Patient-specific seizure detection

We investigated the average firing rate of the output neuron during the ictal and interictal periods to minimize false alarms. This analysis aimed to determine the feasibility of using an optimal universal temporal window to calculate performance metrics. Figure 5 highlights the variability in firing rates between ictal patterns and patients, reflecting differences in seizure dynamics and characteristics. Sensitivity was defined as the proportion of detected seizures in the test set, and delay was calculated as the time from annotated seizure onset to the first 2-second detected interval that met the detection threshold of six spikes. The FAR was measured by counting 2-second intervals with six or more spikes during non-seizure periods and normalizing by the total duration of 20 interictal recording hours per patient. Table I summarizes for each patient the sensitivity, FAR, mean onset delay, and comparison with other works. In 3 out of 5 patients, the SNN achieved zero false alarms and accurately identified all seizure events.

**TABLE 1:** Performance Metrics.

| Patient ID | Sensitivity (%) | | FAR ($\text{h}^{-1}$) | | Delay (s) | |
| --- | --- | --- | --- | --- | --- | --- |
|  | This Work | [28] | This Work | [28] | This Work | [28] |
| ID11 | 100.0 | 100.0 | 0.0 | 0.0 | 14.0 | 19.5 |
| ID12 | 100.0 | 100.0 | 0.0 | 0.0 | 34.4 | 36.3 |
| ID14 | 100.0 | 0.0 | 0.23 | 0.0 | 2.38 | - |
| ID17 | 100.0 | 100.0 | 0.12 | 0.0 | 28.0 | 19.0 |
| ID18 | 100.0 | 75.0 | 0.0 | 0.0 | 15.5 | 25.7 |
| Mean | 100.0 | 75.0 | 0.07 | 0.0 | 18.8 | 25.1 |

**Fig. 5:**
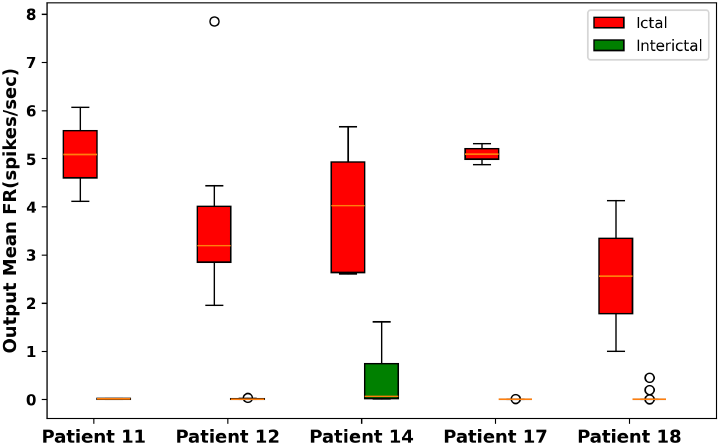
Mean firing rate during ictal and interictal periods for each patient.

## IV. Discussion and Conclusions

We demonstrated that long-term reliable seizure monitoring and detection on a resource-constrained CMOS neuromorphic chip is feasible using population averaging and E-I balance mechanisms. Our proposed solution effectively addressed two common challenges: high FAR and power consumption. The hardware SNN successfully detected all seizure events in 5 patients. Future work aims at the realization of a dedicated ultra-low power ASIC with an Analog Front-End (AFE), an adaptive ADM encoding circuit and the corresponding SNN in one single device. New generations and advancements in specialized devices, particularly with AFE integration and online learning, hold the potential to provide a complete end-to-end solution for low-power implantable and wearable on-edge seizure detection devices. Although the implemented system operates in real time and the chip delivers very low latency, the network currently detects seizure onset activity with a mean delay of 18.8 seconds. Despite this delay, the significant reduction in latency achieved by the system, combined with clinically low FAR, paves the way for closed-loop neuromodulation SoC. Future work will aim to accomplish earlier detection delays to allow the development of closed-loop feedback strategies for real-time seizure suppression and dynamic neuromodulation.

## Data Availability

The original data were derived from the following resources available in the public domain http://ieeg-swez.ethz.ch/.

